## Supplementary figures and images for "Caregiver experiences of an integrative patient-centered digital health application for pediatric type 1 diabetes care: findings from a pilot clinical trial"

### S1 File

Supporting Information 1. Clinic Visit Preparation Form

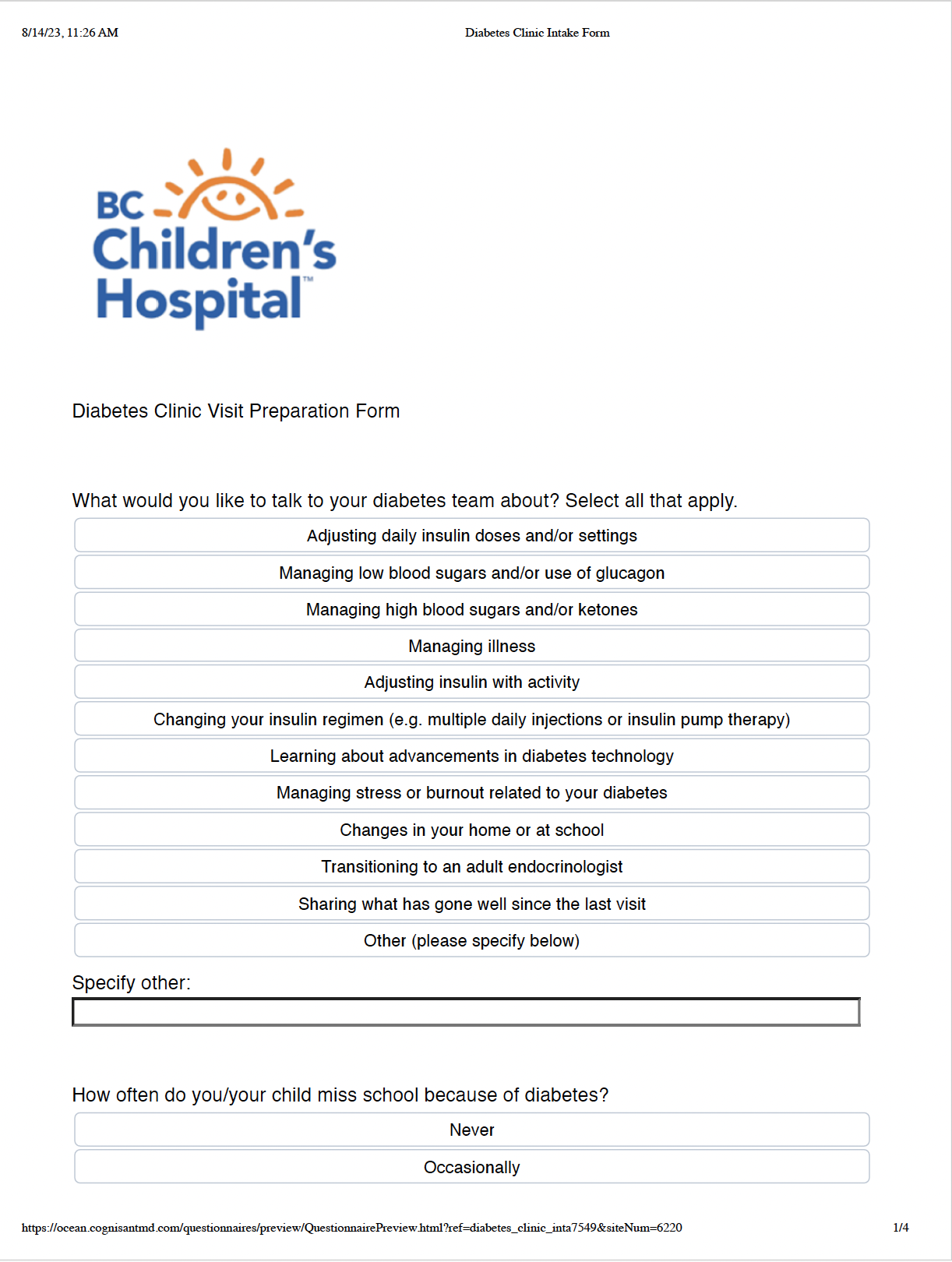

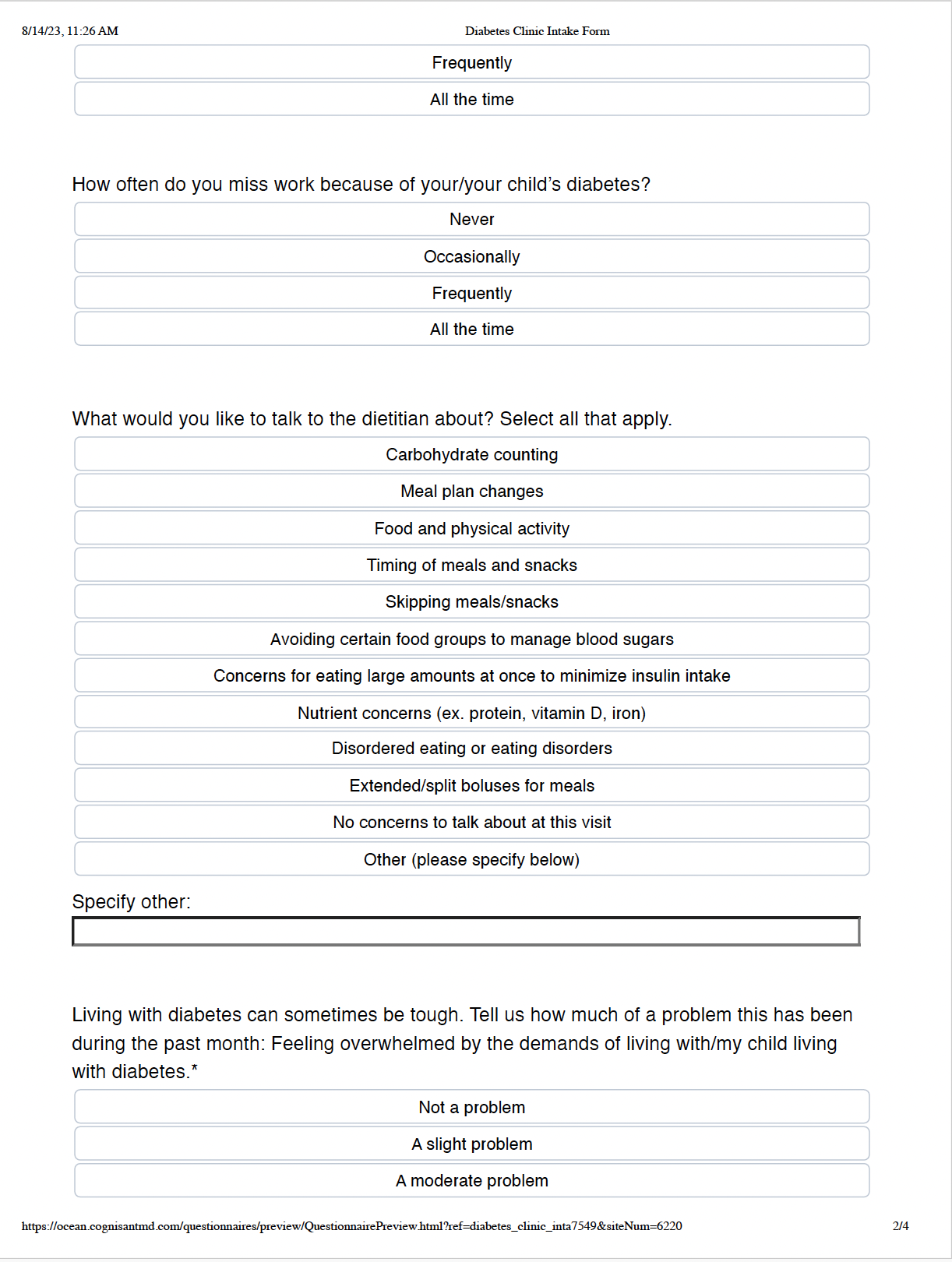

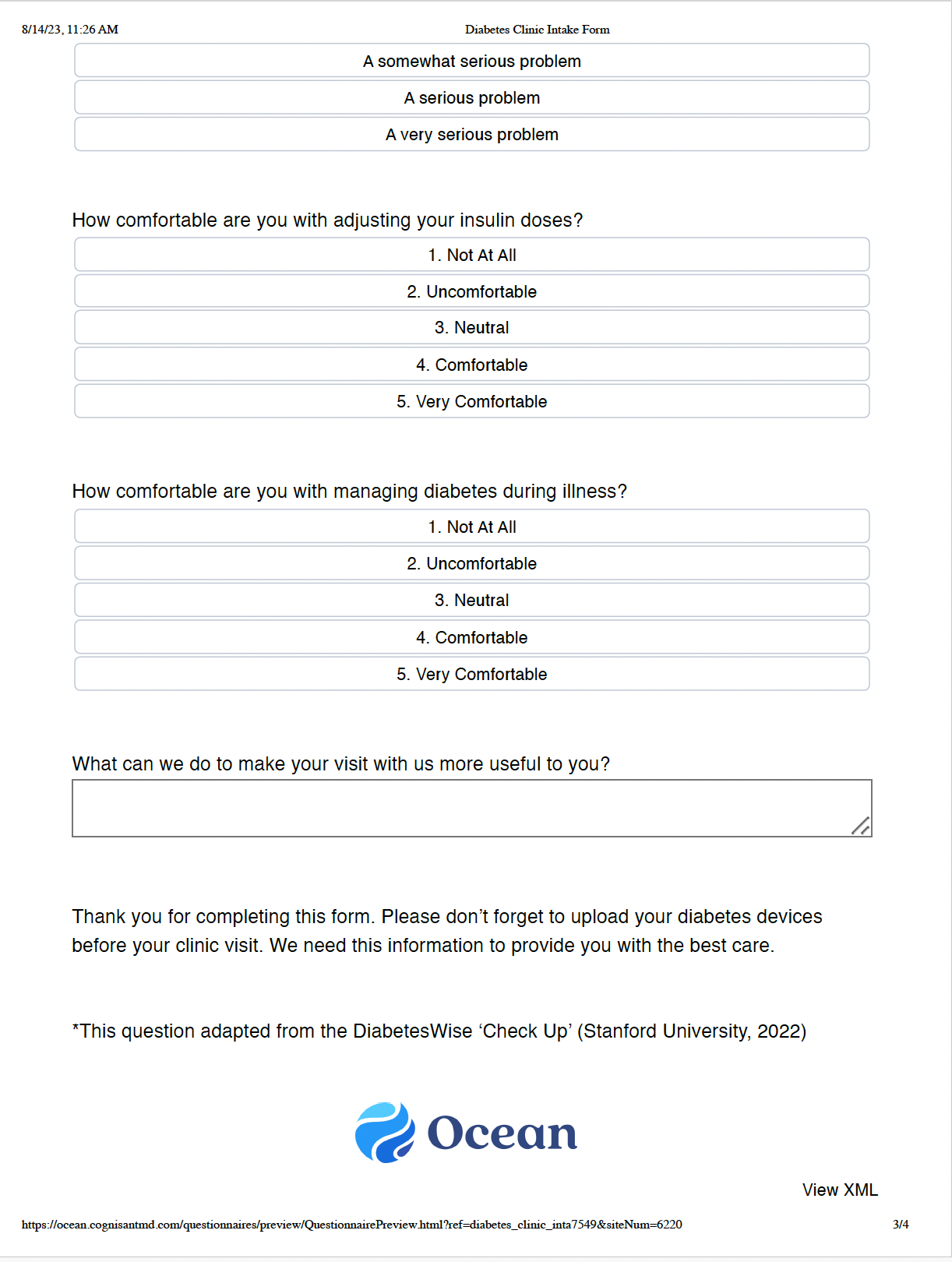

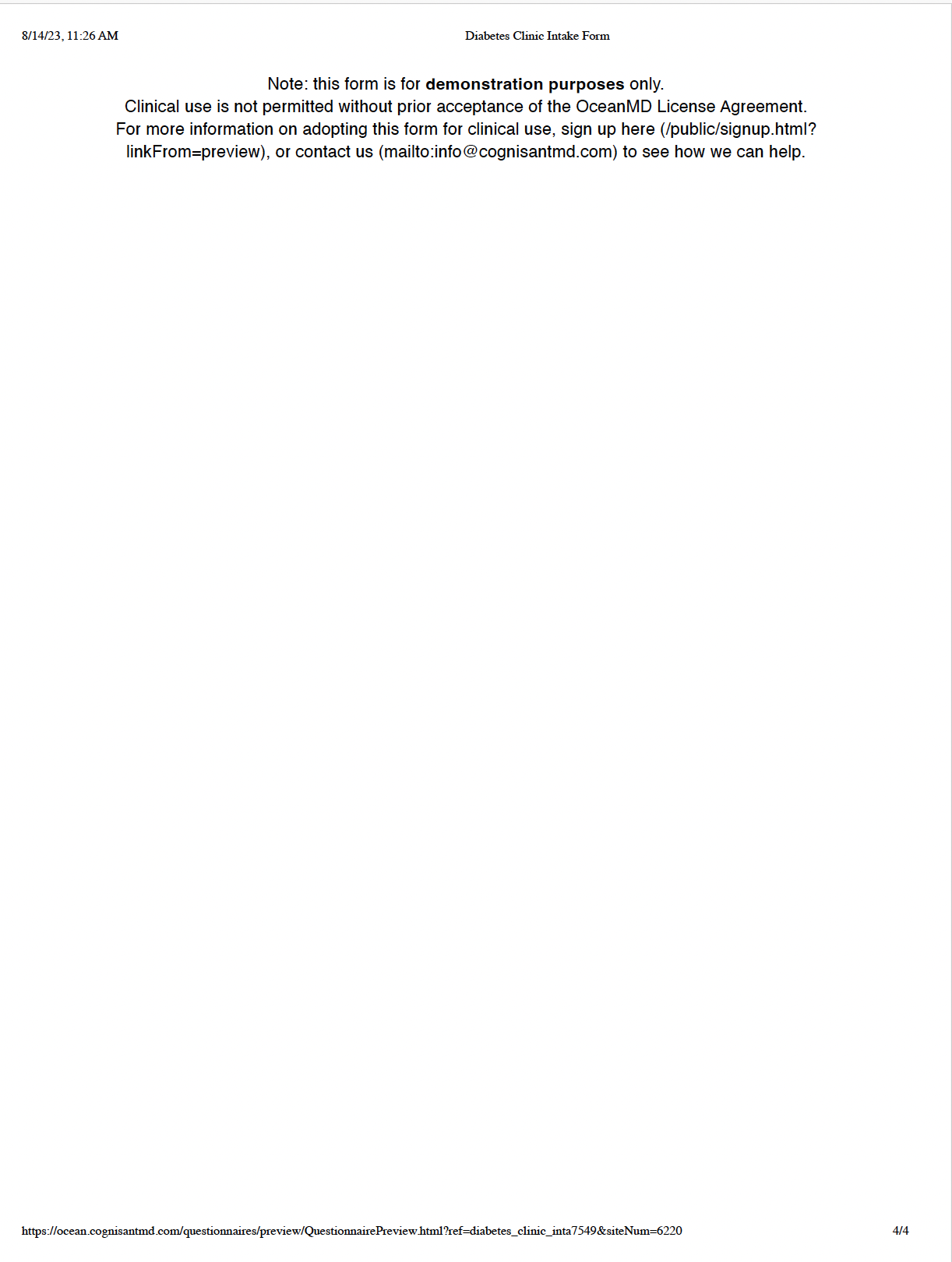

### S4 File

Supporting information 4. Participant Flow

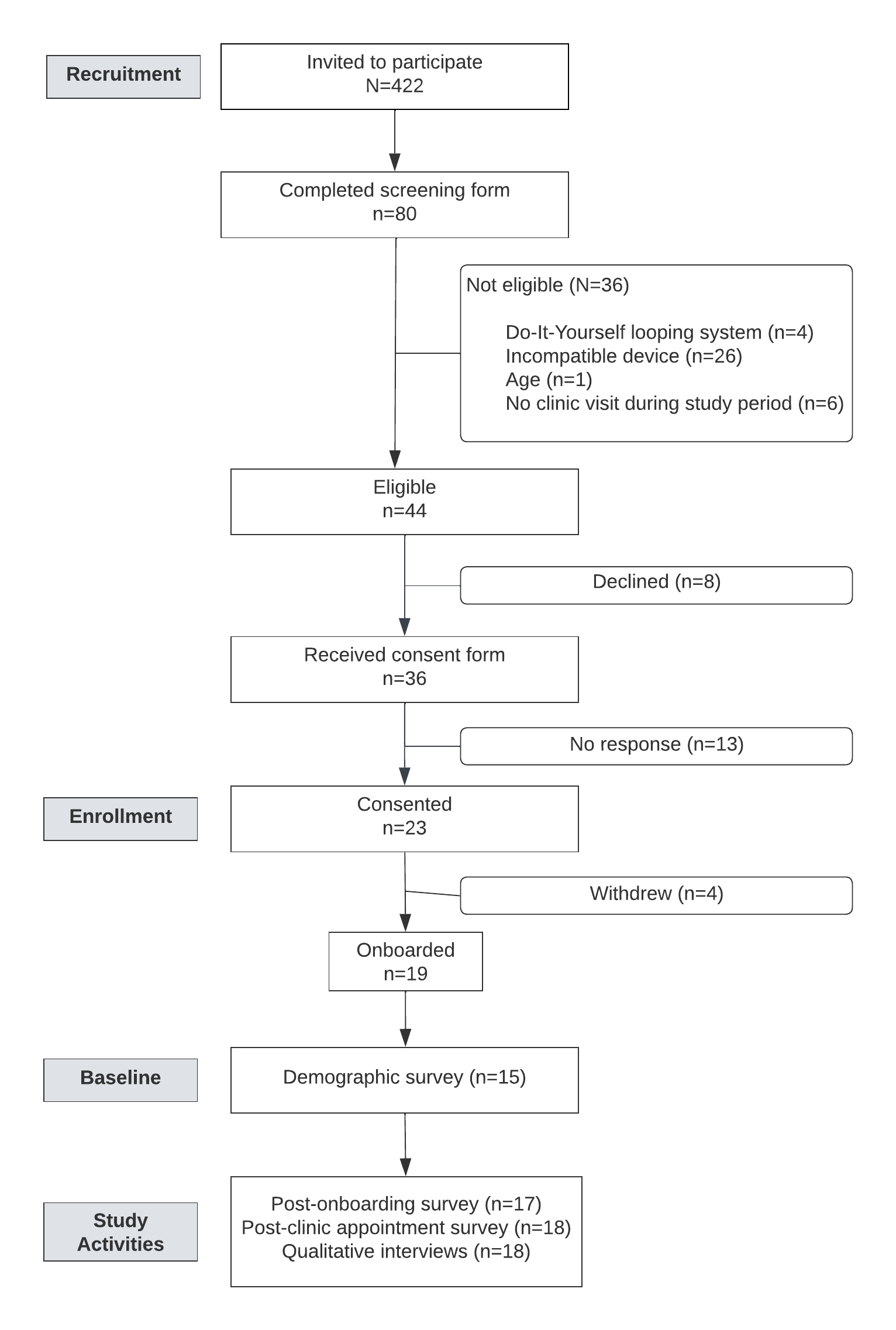
