## Supplementary material for "Caregiver experiences of an integrative patient-centered digital health application for pediatric type 1 diabetes care: findings from a pilot clinical trial": S2 File

Supporting information 2. Post Clinic Appointment Survey

1. Before using Care Hub, how did you usually share your blood glucose data with your healthcare team for your clinic visit? Please select all that apply

- Upload data at clinic visit and healthcare team would print it out
- Upload data at home and healthcare team would print it out before clinic visit
- View data online (e.g. using Dexcom Clarity app) together with healthcare team during my clinic visit
- Other – open text box

2. For this clinic visit, did you use TrustSphere to prepare for your visit?

- Yes
- No

2a. If yes, which functions did you use?

- Intake Form
- Nutrition Assessment
- Viewed glucose levels on Dashboard
- Reviewed and/or completed Tasks

2b. Did these functions help to make you more prepared for your visit?

- Did the _________ help make you more prepared for your visit
  - Yes
  - No

2c. If no, why not? (open text)

3. During this clinic visit, did any of the following members of your healthcare team view your data in Care Hub with you?

- Nurse
  - Yes
  - No
  - Didn’t see them
- Dietician
- Social Worker
- Endocrinologist

4. Do you feel that having Care Hub was helpful to you for this clinic visit?

- Not at all helpful
- Not helpful
- Neutral
- Helpful
- Very helpful

4a. Why did you find Care Hub not helpful? (open text)

4b. Why did you find Care Hub helpful? (open text)

5. Do you feel that your healthcare team was able to make your/your child’s clinic visit and/or their recommendations more personalized because of Care Hub?

- 1. Not at all
- 2. Slightly more
- 3. Somewhat more
- 4. Much more
- 5. Very much more

6. Will you use Care Hub to help you manage your/your child’s diabetes at home following this clinic visit?

- 1. Definitely not
- 2. Probably not
- 3. Possibly
- 4. Probably
- 5. Definitely

6a. Why won’t you use Care Hub at home? (open text)

6b. Why will you use Care Hub at home? (open text)

7. Now that you’ve been using Care Hub for a few months, how would you like your healthcare team to connect with you using Care Hub in between clinic appointments?

- Send me a check-in to find out how things are going related to my/my child's general wellness (i.e. mood, healthy living)
- Send me a check-in about how things are going with a recent change in my/my child's diabetes management (i.e. switch to basal/bolus or technology - pumps, sensor)
- Review my/my child's glucose data and send me recommendations on insulin dose adjustment
- Have a short 15-minutevirtual visit to review my/my child's data on the Careteam dashboard
- Other (open text)

8. Do you use other online tools or apps (besides Care Hub) to view your/your child's glucose trends, insulin and/or carbohydrate data at home?

- Yes, I usually go to other online tools or apps (e.g. Dexcom Clarity, DiaSend, Glooko, FreeStyle Librelink, etc.) to view my glucose data.
- I sometimes view my data in other online tools or apps, and sometimes view in Careteam
- No, I usually go to Care Hub to view my glucose trend data.

8a. Are there other views of your diabetes data that you like to look at that are only available in these other tools or apps? If so, which views? (open text)

9. Do you have any suggestions for how Care Hub could be improved so that it would be more useful for your clinic visits? (open text)
