## Supplementary material for "Caregiver experiences of an integrative patient-centered digital health application for pediatric type 1 diabetes care: findings from a pilot clinical trial": S3 File

Supporting information 3. Caregiver End of Phase Questionnaires

1. Logging in to my app account is easy

- 1. Strongly agree
- 2. Agree
- 3. Undecided
- 4. Disagree
- 5. Strongly disagree

B. What makes it difficult?

- Open text

2. A. Viewing my/my child's clinical information and diabetes data in the Careteam dashboard is easy.

- 1. Strongly agree
- 2. Agree
- 3. Undecided
- 4. Disagree
- 5. Strongly disagree

B. What makes it difficult?

- Open text

3. A. I feel that my/my child's personally identifying information (i.e. name, birthdate) and clinical data (i.e. blood glucose) is secure in this app.

- 1. Strongly agree
- 2. Agree
- 3. Undecided
- 4. Disagree
- 5. Strongly disagree

B. Why do you not feel that your/your child’s information is secure?

- Open text)

4. A. Did you read through the My Data 4 Research consent/assent process to participate in research?

- Yes
- No

B. Why did you not explore this part of the app?

- Open text

C. I found it easy to understand the consent/assent process.

- 1. Strongly agree
- 2. Agree
- 3. Undecided
- 4. Disagree
- 5. Strongly disagree

D. What made it difficult?

- Open text

5. A. I would recommend use of this app to other families with children living with T1D.

- 1. Strongly agree
- 2. Agree
- 3. Undecided
- 4. Disagree
- 5. Strongly disagree

B. Why would you not want to recommend the app?

- Open text

6. Are there any additional comments you would like to add regarding your experience with this app?
   - Open text
