## Supplementary material for "Caregiver experiences of an integrative patient-centered digital health application for pediatric type 1 diabetes care: findings from a pilot clinical trial": S5 File

Supporting information 5. Qualitative Interview Themes, Subthemes and Quotations

**Theme 1. Convenient Connection**

Convenience and Handiness

"So it’s nice to have it mobile...in our hand, and we used to have to print stuff out and take all these papers with us to Children’s... We can go 'oh, let’s look at this'...So there’s some convenience to that its portability." (P2)

Here’s some reading when...I’m at the hockey rink or at the dance studio all my kids are there; that’s a super amazing thing to have because you’re not trying to go find it, it’s already there for you.” (P6)

“I think apps are the way to go for sure…everybody wants something at the touch your finger. So the app, the fact that you can have it on like an iPhone…it’s so handy, like that’s what I really liked is the option of being able to do this stuff through an app is way better than trying to email in or any of that stuff, because you always have access to your phone…The app, it’s just everything all in one place, you don’t have to go searching for something.” (P12)

“It has the potential to be a really useful on hand tool with readily available information. And it’s easy if you have 5 minutes just to quickly look at perhaps [child’s], you know, the data or just the prompts…rather than everything getting lost in the hundreds of junk emails that you get every day.” (P4)

“If we’re at home... we have like her insulin needs ...so I like being able to like go to the hospital and not have to bring that with me. I just have it all in the app... Like her sliding scale numbers and what her insulin doses are... it’s nice to not have to even think about [it]...like it’s just there right”. (P1)

“The features that I liked were just having all the resources about the diabetes together. Like I have my binder with me when I go on trips, but for example, right now, I’m in town and I don’t have my binder, I have my phone. And it was nice knowing that it was all in one file but like on your app.” (P17)

Empowered

“When we look at the 30 day and the 90 day...I like to be able to see that and...your glucose levels and your time in range and all that type of thing that’s neat to see...we don’t look at this data in any way, shape or form as a sort of report card...when we analyze the graph and look at them and go...'in the morning you’re doing this, maybe we should tweak that it'. That truly is very helpful and it helps us understand.” (P2)

“The motivators are the ease of use and looking at a way to engage [child] in the process. I feel sort of having an app might make it easier for him to look up things and be comfortable looking up things. Because he’s pretty good and, you know, he sort of self-manages...but perhaps...that’s a tool for him that he could easily look at if he needed to check something.” (P4)

“I think that for me, it was really helpful seeing that she was high a lot of the time, so I think it improve[d] my treatment, overall.” (P8)

When the doctor input his recommendations after the visit, it’s nice that that will be in there, I believe forever...But then looking back we can see...what the suggested basal rate changes were, or the change for the insulin to carb ratio, or the correction factor, that sort of thing. So it’s helpful to have that all in there. (P11)

Connected

“I think…[it] makes the communication and everything a little bit more accessible with the endocrine team. So... if you want to call it a bridge like it kind of bridges between home care and the hospital care.” (P1)

“The only part I had found useful was just communicating with the doctors, like when they had information for me to forms or what not they wanted me to fill out. They. I could see that they had messages for me there. So it was a good way to communicate back and forth.” (P17)

“It was beneficial, for sure because it was...personal... It was a message that [my child’s] doctor actually entered to follow up after the appointment and touch base...it was a friendly note with the recommendations...a little bit of a better connection because it was like he took the time to...send us the information. Instead of, at the previous appointments, I would have to make sure to write it down and remember it.” (P11)

“Really just that main thing that like really preparing me for the clinic appointment. Because, like as I said, like, I probably would have communicated with the care team if that option was available through the app...But prepping for the appointment, I felt good about it. I felt like there was a communication factor there, even if it wasn't necessarily like it was just more questionnaire.” (P9)

Prepared

“The most beneficial use I’ve had with it is being prepared for my appointments with the doctors, the team at the diabetes clinic. It reminds me to take my measurements and submit information and it kind of focuses our questions my questions that I may have.” (P17)

“What I liked the most was how it did actually help me prepare better than I prepared in the past for her clinic appointment.” (P9)

They sent those forms to fill out beforehand. I did like that... So that really definitely helped us prepare for the meeting and to focus our questions for the team.” (P13)

“The thing that stands out the most, I guess, would be in preparation for my child's endocrine appointment they were able to... I can't remember what the word is…a pre screen? They were able to do like the questionnaire before the actual appointment, which was helpful…Usually they send you a letter, and it's like by mail, and you have to write out your blood sugars and all these things. It was nice to be able to do that digitally on the app. (P1)

“The diabetes clinic team requested in the app that I upload his pump data before the appointment, and that was helpful to have that in there. And they did it, and they could just access it through the app, that's a really beneficial thing, to communicate that way.” (P11)

In the past when I filled in the paper and brought it to the appointment it was. It’s probably not enough time for the nurses or dietitians, whoever is going to see us that time...for them to grab that paper and review it. Once, you know, we come to the office and then our appointment is right away, so it’s better if they can see it on the app and they can review it before we come. (P11)

I used it in preparation for the appointment...So it kind of helped me prepare a little bit better for the appointment, because I guess it made me have to sit and think for a second on like questions that I had for my clinic appointment. So those prompts before the appointment were actually helpful and I used it. (P9)

**Theme 2. Usage Drives Value**

Clinical Team Usage

“I’m willing to use it… to communicate with the care team, to prepare for clinic appointment. Yeah, like If the care team were to give me like some things to do within the app, I would definitely be happy to use it that way.” (P9)

“For me to continue seeing a benefit from it in the future...would be more integration with our healthcare team, more prompts...For me the big cherry would be that integration with Children’s Hospital and have that push format...to create more engagement. (P5)”

“It has been a very valuable tool in terms of that data…I think it does a very good job of collecting and collating all the data that comes from all these different types of sources when it comes to the care of diabetes...It’s our hope, my hope, that that data doesn’t just sit there and it gets used...There’s a responsibility for us to use it, which we do, but also a responsibility for the team at Children’s to also have that data at their fingertips to be able to provide good recommendations.” (P10)

“I’m able to enter information and also the doctor can send notes or information in the app as well. And so after our appointment, the doctor entered the recommendations into the app, and I was easily able to access that, so that was excellent; very helpful.” (P11)

“The doctors have loaded some things up…the care plan, after we’ve had a visit, they’ll load information on there and if I need to kind of look at it for reference, because sometimes the appointments happen really quickly, I have that as a reference which has been really helpful to use.” (P15)

“The other thing I would say that is a benefit would be like just communication between the doctor or the nurse and us. Like I know they're able to upload forms and I imagine…they can upload other relative information or share information via the hub. So, I thought that was very helpful rather than having to wait for your appointment…It's like a central place to be able to act rather than via email or whatever.” (P1)

“I think what would be more helpful to me is if on the... healthcare team side that they were pushing the information to me more...here’s your next appointment confirm it through the app. Here’s some reminders that you need to get this blood work done by this time...I would love that information to be pushed to me more instead of me inputting it...because that would be more helpful...Reminders on the blood work...Celiac...She had her A1C done at our last appointment and I have a paper copy of that but I don’t have that anywhere in my Careteam or attached to our last visit.” (P5)

“I would be happy to see if the doctors or nurses put recommendations in there and if that’s stored in there that would be excellent.” (P11)

Central Repository or Hub

“I think...having everything in...as much as possible in one app is kind of nice, just so you don’t have to have so many on your phone. I think that it consolidates a lot of resources.” (P8)

“For us, it’s not my daughter who’s taking care of her diabetes, it is really a team. It’s myself, my husband, my parents, her endo team, and we can all use that as kind of a you know a hub.” (P1)

“Just being able to for your care team to like access all of the numbers and any information and just having that connection that way." (P18).

“I would recommend it because of having all of the information stored in one place. So like that is really positive and helpful for just getting an overall picture of your child's like diabetes situation, if you will.” (P9)

“It made it [sharing information] easier because if they can just see what’s going on, if we’re all connected more easily, then the team can see it easier and we can see it easier.” (P14)

“Yeah, so it has been positive that way, as a tool to share... information with the doctor.” (P17)

“I think the sharing aspects…like if you’re all on the same team like my son and I are together…you can chat about, ‘what do you think about this?’ And my child could look at his and go, ‘Well, I think this’. Just some sort of communication piece between people on the same care team inside, like especially in the dashboard, when you’re looking at the graphing and stuff.” (P2)

“I like the idea of being able to input kind of your extended family and things like that in there and that information…I think that it is great to be able to have, you know, this information in one place. (P5)”

“I think...having everything in...as much as possible in one app is kind of nice, just so you don’t have to have so many on your phone. I think that it consolidates a lot of resources. And again, like I mentioned, the showing the data over time, showing like to show the trends is important. I think having the forms in there is handy as well. (P8)”

**Theme 3. Hope and Future Benefit**

Hopefulness

“The app to me is, where it’s at right now, is phenomenal in what it does so far. I see huge potential in it for the future.” (P6)

“It was exciting to be a part of it, because it is useful and beneficial to families living with diabetes...and hopefully it sticks and it will be something that will be used.” (P11)

“So I do hope that they do use it, and that there’s good training on it, and that they will embrace it as a way to communicate because we’re all for these type of things that make life easier. And [it] can be a great communication tool and helping to, you know, have him have a more productive and healthy life, ultimately.”(P10)

Future Benefit with Bi-directional Communication

“I think it would be helpful is if there was [a] live chat option that you could pop up and say…talk back and forth with me, or help me troubleshoot what’s going on, even if it’s like there’s a slight delay in their response...People I think, and myself included, are hesitant to want to call Children’s but the reality is…having someone on the other end of that to be able to quickly like text... having that on the app would be helpful just even if...you get a response within 24 hours. Because I think there is that natural hesitancy...to call the doctors if it’s not an emergency. I know that is the case for myself... it's the absolute last resort for me to call Children’s Hospital because it’s not what we do.” (P15)

“I may have like one one-off kind of question that would be helpful for me to know but then if it was just like one one-off question I might not be as motivated to write an email and ask. But if I had the ability to contact, just like almost like a text message conversation through the app then I would totally be more on point with getting questions that kind of pop up in the moment answered. Which overall, I think that getting these things figured out no matter how small it is, it’s kind of like giving me the opportunity to understand the whole picture of, like her diabetes care a lot better”. (P9)

“Maybe if there were like a notification of, I don’t know, like over the last 7 days your, they’ve been high at such and such time or something like that. Like some kind of notification if there is an alert that maybe spotted particularly troubling trends, that might be helpful.” (P8)

“I think it could be utilized more. I think that they could be, you know communicating, especially with some of the little ones where, you know, we have to make frequent changes more with their insulin doses. To communicate, you know, instead of like every 3 months maybe to talk to like somebody via the app, a nurse or something, to say, hey, you know, maybe make a change here or a change there. I feel like that could be more beneficial. I mean, these little kids they tend to need changes so frequently as they are growing and changing, and as a parent, I feel like I'm still scared to make those changes without kind of the guidance and reassurance from a nurse”. (P14)

“The thing that I like the best about it is that I feel like it’s open communication and like current and accessible information between us and our Endo team. So I love that you know that we can both look at the same thing essentially...I think the things that we are using such as like the forms and the communication, the chat between the doctors, and then just being able to like leave notes...instantly, I think that’s really beneficial”. (P1)

"I’d love to see it developed more as a good hub of communication where they can … make suggestions or we can use that data to turn into something more useful to help better his care”.

So it would be, to have a two-way street in terms of the care that we’re giving to our son and that they can give to him, in terms of like, hey, I’ve noticed this over the last three weeks this has been happening, here’s what you should do to bring those numbers down, or bring them up, or whatever. So I’d love to see it developed more as a good hub of communication where they can action, where they can make suggestions, or we can use that data to turn into something more useful to help better his care. (P10)

I don’t mind emailing back and forth, but it’s almost like the difference between email and text message, where...It’s not as like choppy. It’s not as disconnected where it’s like one email back to you, one email back to me, where it’s like it’s just a continuous conversation is what it feels like…I think that I would inquire about different things a little more often than I do if that option were there, just because I feel like when it comes to drafting an email or writing up an email, for me, it feels like it takes a lot more effort out of me than to just like for example, if I were just to send somebody a text message that’s a little bit more informal. So I don’t feel like I need to like put as much thought into like how I’m structuring my sentences, or whatever. So I feel that I would communicate a lot more with the team if that option were there, or that part of the app was available to use. Yeah, those are the those are the things that I like to the most. (P9)

I think I would use this app in order to get in touch with the clinic, the nurses the diabetes clinic nurses, because it is very difficult to call them and get an actual person to talk to right away. So, for me, it would be more how can I gain access to them faster…how can I get better like faster information that I need when I want to change part of his treatment? And I would hope that the app would do that, since we’re always looking at it. And I would hope that someone would be able to look at it as well, since we did give permission to follow him as well. (P13)
